# Implementation and utilization of real-time prescription benefit tools across three large academic health systems

**DOI:** 10.1101/2024.12.27.24318670

**Authors:** Muhammad Ali, Jeremy I. Schwartz, Jiangxia Wang, Jing Liu, Niteesh Potu, William Rottier, Peter Dziedzic, Erika Smith, Tanvi Mehta, Nitu Kashyap, Michael J. Fliotsos, Jing Luo, Jeremy Epstein, Jodi B. Segal, Harold Lehmann, Bradley H. Crotty, Fasika A. Woreta

## Abstract

**Background:** Real-time prescription benefit (RTPB) tools provide clinicians an out-of-pocket cost (OOPC) estimate at the time of prescribing and may help them select medications with lower OOPC for their patients.

**Objective:** To evaluate how RTPB tools altered medication orders during clinical encounters and assess patient and clinician characteristics associated with the display of RTPB tool and alternative selection.

**Design:** Retrospective cohort study

**Setting:** Outpatient encounters at three academic medical centers

**Participants:** Patients and clinicians

**Measurements:** Patient characteristics, clinician characteristics and medication alternatives suggested by RTPB tool were compared across the three sites.

**Results:** From April 2019 to October 2021, 6,562,442 patient encounters occurred between 3,624 clinicians and 1,261,551 unique patients. Medications were prescribed in 2,152,772 (32.8%) encounters, of which RTPB tool retrieved and displayed alternatives in 968,811 (45.0%) encounters. The clinician selected an alternative in 68,731/968,811 (7.1%) of the encounters during which 89,050 medications were prescribed. The unit cost of alternative medications remained the same for most orders (n=41,212; 58.4%), while 18,629 (26.4%) had lower cost and 10,728 (15.2%) alternate orders had higher cost. Clinicians selected a different pharmacy among 39,634 (44.5%) and a different pharmacy type (mail vs retail) among 7,508 (12.7%) of the alternate medication orders, of which most were to mail order 4,680 (62.3%).

**Limitations:** We could not assess the role of pharmacy benefit manager coverage, cash-based alternative pricing, and impact of prior authorizations which may be assessed during future investigations.

**Conclusion:** Alternate prescribing after implementation of RTPB tools was low across the three institutions. Unit cost of the medication did not often change. Most changes reflected pharmacy choice. Further studies are needed to assess the barriers in adoption of RTPB tools.

## INTRODUCTION

Cost-related nonadherence, largely driven by high out-of-pocket costs (OOPC), is one of the most common causes of medication nonadherence in the US.(1) Accurate OOPC information could help prescribers select medications with lower OOPC for their patients.(2) Starting January 2021, Medicare Part D plans required all electronic health records (EHR) to incorporate a real-time prescription benefit (RTPB) tool, to enable price transparency and cost saving alternatives for patients.(3–5)

RTPB tools, embedded in EHRs, are designed to increase price transparency at the point of prescribing.(6) An RTPB query is generated at the time of medication order placement within the EHR via contracted RTPB vendors and is sent to pharmacy benefits managers (PBMs). PBMs then cross-reference patients’ health plan details with PBM’s insurance and prescription coverage and return OOPC for patients with prescription coverage. Resultantly, the EHR-embedded RTPB tool retrieves and provides cost estimates based on patients’ specific insurance plans and options for lower cost intraclass medication alternatives or pharmacy alternatives when applicable.

The use of an RTPB tool in a cluster randomized trial showed that while only 4.2% of medication orders displayed an eligible alternative, there was an 11.2% reduction in OOPC for these orders and a 38.9% reduction in the OOPC of high-cost drugs.(7) Bhardwaj et al. found that the use of an RTPB tool was associated with an increased prescription fill rate at pharmacies.(8) Sinaiko and colleagues found that clinicians changed medications more often when potential cost savings were ≥ $20 and for certain drug classes like asthma and chronic obstructive pulmonary disease medications, antihyperglycemic agents, and cardiovascular medications.(9)

The real-world effects of these tools bear further investigation in a multi-institution approach, as the impact of RTPB tools may depend on coverage of contracted PBMs, site-specific implementation choices, and patient and clinician factors.(10) Further data are needed to determine how and whether these tools alter treatment plans or steer dispensing towards particular pharmacies or mail-order services, which can have additional policy or value considerations. To address these questions, we used a federated query scheme across three academic medical centers to assess patient and clinician characteristics that were independently associated with display of alternatives by the RTPB tool and subsequent selection of an alternative by the clinician.

## METHODS

### Data

We extracted outpatient encounter data from the EHR of three large US academic health systems between April 2019 to October 2021. All three sites used Epic^®^ (Epic Systems, Verona, WI) as the electronic health record. We extracted patient characteristics including age, gender, race, ethnicity, health insurance type, and total number of medications at the time of encounter. We also extracted clinician characteristics including sex, type (attending physician, nurse practitioner, resident physician, etc.), specialty, rate of medication prescribing (number of encounters with medications ordered out of the total number of encounters) and the number of times RTPB displayed options. Medication characteristics included therapeutic class of medication, total cost, unit cost, quantity, pharmacy type (e.g. mail, retail), pharmacy (e.g. Walgreens, CVS, etc.) and medication name.

### Eligible Visits

We included all RTPB eligible in-person or virtual visits conducted in an outpatient setting across all clinical specialties. Visits with clinicians with fewer than 50 encounters with medication orders across the observation period were excluded. We excluded requests for refills and telephone encounters because these medication orders are often placed by other members of the care team who do not make decisions on alternatives and refills requests indicate medication adherence. The study protocol was approved by the Johns Hopkins University School of Medicine Institutional Review Board and all study activities adhered to the tenets of the Declaration of Helsinki.

### Study Outcomes

The primary outcome of our study was the use of the RTPB tool. The use of the tool was considered complete when a medication order triggered the RTPB tool, the OOPC estimates were retrieved based on the patient’s insurance plan, and options for lower cost intraclass medicines or pharmacy alternatives were displayed to the clinician in real-time.

Our secondary outcome was the clinician’s selection of an alternative option as suggested by the tool. To characterize the alternatives, we recorded the therapeutic class of medication, total cost variance, unit cost variance, change in quantity, change in pharmacy type (e.g. mail, retail), change in pharmacy (e.g. Walgreens, CVS, etc.) and change in medication. Change in medication was further stratified by whether it was a change in formulation, strength, quantity, or a combination of these. Medication cost was classified into quartiles for the multivariable model. Quartiles were defined as 1st Quartile: < $0.94, 2nd Quartile: $0.95 - 4.28, 3rd Quartile: $4.29 - 10.51), and 4th Quartile: > $10.52.

### Statistical Analysis

After pooling data across the three sites, we compared characteristics of clinicians who selected an alternative medication at least once vs those who never selected an alternative. Chi-squared tests were used to test for differences in categorical variables, and Students’ t-test and Mann-Whitney U tests for continuous variables.

Using encounter level data, we used multivariable logistic regression to test which patient and clinician characteristics were independently associated with the display of alternatives by the RTPB tool. Using medication-order level data, we also used multivariable logistic regression to test which patient and clinician characteristics were independently associated with selection of the alternative by the clinician. To reduce the complexity of the multilevel structure of the data (i.e., multiple encounters per patient across different clinicians) and computational burden of running the complex models on the entire dataset, a multiple outputation approach was used by running population-average logistic regression models on reduced samples by bootstrapping i.e. randomly sampling one encounter per patient from 5% of the entire sample. This procedure was repeated 1000 times to estimate the marginal risk ratios (RR) with corresponding 95% confidence intervals (CI) and p values.(11) The reduced samples streamlined computation and avoided clustering from same patients going to different clinicians so that the clustering is only at the clinician level. All statistical analyses were performed using Stata Version 17.0 (College Station, Texas, USA) and statistical significance was defined as p<0.001.

## RESULTS

All three institutions utilized Epic Systems’ (Verona, WI) electronic health record although they made different RTPB tool implementation choices. Surescripts (Arlington, VA) and RxRevu (Denver, CO) were the two RTPB vendors utilized at the study institutions. Sites A and B had contracted one RTPB vendor each whereas site C had a contract with both vendors. The tool displayed automatically for eligible orders at sites A and C, whereas clinicians had to manually request estimates at site B. Clinicians had the option to opt out of the tool at all three sites. There was no cost differential threshold at sites A and B whereas site C had a cost differential threshold of $3/month or $0.10/day, below which the alert would not display.

We had data from 6,562,442 encounters of 1,261,551 unique patients to 3,624 clinicians across the three institutions. Most encounters were in-person visits (n= 5,522,389; 84.2%), followed by virtual visits (n=841,698; 12.8%), and procedure visits (n=198,355; 3.0%). The mean age of patients was 53.0 (±18.6) years with most patients being female (n=744,063; 59.0%). The patients were 69.8% White (n=880,246), 14.6% Black (n=184,531), 3.4% Asian (n=43,367), and 7.9% reported Hispanic ethnicity (n=99,878). Most patients had commercial insurance (n=705,279; 55.9%), followed by Medicare (n=371,184; 29.4%), Medicaid (n=118,455; 9.4%) and veteran/ military insurance (n=34,423; 2.7%). **(Table 1 and 2)**

**Table 1:** Characteristics of outpatient encounters ^a^ at three academic medical centers from April 2019 to October 2021.

|  | Site A | Site B | Site C | Total | p-value |
| --- | --- | --- | --- | --- | --- |
| **Total<sup>a</sup>, n** |  |  |  |  |  |
| **Total number of outpatient encounters** | 996,138 | 3,322,787 | 2,243,517 | 6,562,442 |  |
| **Encounter type** |  |  |  |  | <0.001 |
| In-Person Visit | 742,059 (74.5%) | 2,886,145 (86.9%) | 1,894,185 (84.4%) | 5,522,389 (84.2%) |  |
| Virtual Visit | 239,696 (24.1%) | 309,896 (9.3%) | 292,106 (13.0%) | 841,698 (12.8%) |  |
| Procedure Visit | 14,383 (1.4%) | 126,746 (3.8%) | 57,226 (2.6%) | 198,355 (3.0%) |  |
| **Total number of unique clinicians** | 791 | 1,769 | 1,064 | 3,624 |  |
| **Total number of unique patients** | 285,034 | 616,193 | 360,324 | 1,261,551 |  |
| **Total number of outpatient encounters with medication order placed** | 420,767 (42.2%) | 988,367 (29.8%) | 743,638 (33.2%) | 2,152,772 (32.8%) | <0.001 |
| **Total number of outpatient encounters with medication order placed and medication estimate generated** | 211,009 (50.2%) | 302,165 (30.6%) | 455,637 (61.3%) | 968,811 (45.0%) | <0.001 |
| **Total number of outpatient encounters with medication order placed and medication estimate generated, and clinician selected alternative** | 14,941 (7.1%) | 1,408 (0.47%) | 52,382 (11.5%) | 68,731 (7.1%) | <0.001 |

<sup>a</sup> Encounters included face-to-face encounter types (both in-person and virtual). Encounters primarily with ancillary staff such as nurses and pharmacists and those providers with less than 50 encounters with medication orders were excluded. Refill, telephone, and erroneous encounters were also excluded.

**Table 2:** Characteristics of all *unique* patients from outpatient encounters ^a^ at the three academic medical centers from April 2019 to October 2021.

| Characteristic | Site A | Site B | Site C | Total | p-value |
| --- | --- | --- | --- | --- | --- |
| **Total, n** | 285,034 | 616,193 | 360,324 | 1,261,551 |  |
| **Age, mean (SD)** | 51.9 (17.9) | 54.2 (18.6) | 52.0 (18.9) | 53.0 (18.6) | <0.001 |
| **Patient's gender** |  |  |  |  |  |
| Female | 171,485 (60.2%) | 365,172 (59.3%) | 207,406 (57.6%) | 744,063 (59.0%) | <0.001 |
| Male | 113,515 (39.8%) | 251,011 (40.7%) | 152,906 (42.4%) | 517,432 (41.0%) |  |
| Other (Null, Unknown, or Non-binary) | 34 (<1%) | 10 (<1%) | 12 (<1%) | 56 (<1%) |  |
| **Race and ethnicity** |  |  |  |  |  |
| White | 161,600 (56.7%) | 433,644 (70.4%) | 285,002 (79.1%) | 880,246 (69.8%) | <0.001 |
| Black | 75,468 (26.5%) | 64,492 (10.5%) | 44,571 (12.4%) | 184,531 (14.6%) |  |
| Hispanic | 17,043 (6.0%) | 67,456 (10.9%) | 15,379 (4.3%) | 99,878 (7.9%) |  |
| Asian | 18,755 (6.6%) | 15,810 (2.6%) | 8,802 (2.4%) | 43,367 (3.4%) |  |
| Others | 10,789 (3.8%) | 21,001 (3.4%) | 5,395 (1.5%) | 37,185 (2.9%) |  |
| Unknown | 1,379 (0.5%) | 13,790 (2.2%) | 1,175 (0.3%) | 16,344 (1.3%) |  |
| **Insurance** |  |  |  |  |  |
| Commercial | 181,234 (63.6%) | 327,550 (53.2%) | 196,495 (54.5%) | 705,279 (55.9%) | <0.001 |
| Medicare | 66,655 (23.4%) | 190,619 (30.9%) | 113,910 (31.6%) | 371,184 (29.4%) |  |
| Medicaid | 1,858 (0.7%) | 81,415 (13.2%) | 35,182 (9.8%) | 118,455 (9.4%) |  |
| Veteran/ Military Insurance | 26,914 (9.4%) | 4,567 (0.7%) | 2,942 (0.8%) | 34,423 (2.7%) |  |
| Null | 3,987 (1.4%) | 5,900 (1.0%) | 8,080 (2.2%) | 17,967 (1.4%) |  |
| Other/ Self Pay | 4,386 (1.5%) | 6,142 (1.0%) | 3,715 (1.0%) | 14,243 (1.1%) |  |
| **Total number of medications at time of encounter, median (IQR)** | 3.0 (0.0, 7.0) | 3.0 (0.0, 7.0) | 4.0 (1.0, 8.0) | 3.0 (0.0, 8.0) | <0.001 |

<sup>a</sup> Encounters included face-to-face encounter types (both in-person and virtual). Encounters primarily with ancillary staff such as nurses and pharmacists and those providers with less than 50 encounters with medication orders were excluded. Refill, telephone, and erroneous encounters were also excluded.

Medications were prescribed in 2,152,772 (32.8%) encounters, of which the RTPB tool triggered and displayed alternatives in 968,811 (45.0%) encounters. Site B, which required clinician action to display the tool, had the lowest percentage of encounters (n=302,165; 30.6%) during which the RTPB tool displayed alternatives, compared to sites A (n= 211,009; 50.2%) and C (n=455,637, 61.3%; p<0.001).

The clinician selected an alternative in 68,731 of 968,811 (7.1%) encounters with a total of 89,050 medication orders. Site C, which had a cost differential threshold for display of the tool, had a higher percentage of encounters where an alternative was selected (n=52,382, 11.5%) among those encounters where the tool displayed alternatives relative to sites A (n=14,941, 7.1%) and B (n=1,408, 0.47%; p<0.001).

Among the 3,621 clinicians for whom the RTPB tool displayed options, 2,402 (66.3%) used the tool to select an alternative medication at least once. Clinicians who selected an alternative medicine at least once had higher median number of encounters during which RTPB tool displayed options (187.0; Interquartile range [IQR]: 71.0- 535.0 compared to those who did not select an alternative: 46.0; IQR: 22.0-114.0; p<0.001). **(Table 3)**

**Table 3:** Characteristics of clinicians who never selected RTPB alternative versus who selected the alternative at least once from April 2019 to October 2021.

| Characteristic | Never Selected Alternative | Selected Alternative | p-value |
| --- | --- | --- | --- |
| **Total, n** | 1,219 | 2,402 |  |
| **Sex** |  |  | 0.001 |
| Female | 689 (56.5%) | 1,364 (56.8%) |  |
| Male | 508 (41.7%) | 1,025 (42.7%) |  |
| Unknown | 22 (1.8%) | 13 (0.5%) |  |
| **Type** |  |  | <0.001 |
| Attending physician | 669 (54.9%) | 1,620 (67.4%) |  |
| Nurse practitioner | 236 (19.4%) | 368 (15.3%) |  |
| Resident physician | 137 (11.2%) | 141 (5.9%) |  |
| Physician assistant | 89 (7.3%) | 145 (6.0%) |  |
| Fellow physician | 37 (3.0%) | 38 (1.6%) |  |
| Pharmacist | 17 (1.4%) | 21 (0.9%) |  |
| Other <sup>a</sup> | 34 (2.8%) | 69 (2.9%) |  |
| **Specialty** |  |  | <0.001 |
| Medicine, Specialty | 290 (24.0%) | 719 (29.9%) |  |
| Medicine, General | 218 (18.0%) | 522 (21.7%) |  |
| Obstetrics & Gynecology | 85 (7.0%) | 198 (8.2%) |  |
| Family Medicine | 68 (5.6%) | 260 (10.8%) |  |
| Ophthalmology | 44 (3.6%) | 108 (4.5%) |  |
| Surgery, Specialty | 127 (10.5%) | 138 (5.8%) |  |
| Other <sup>b</sup> | 387 (31.2%) | 457 (19.0%) |  |
| **Rate of medication prescribing, mean (SD)** | 33.5 (21.2) | 34.5 (17.9) | 0.16 |
| **Number of times RTPB displayed options, median (IQR)** | 46.0 (22.0, 114.0) | 187.0 (71.0, 535.0) | <0.001 |
| **% encounters with RTPB displaying options, mean (SD)** | 34.8 (24.4) | 47.3 (19.9) | <0.001 |

<sup>a</sup> Others include chiropractor, midwife, medical assistant, technologist and social worker.
<sup>b</sup> Others include radiology, emergency medicine, anesthesiology, dentistry, dermatology, general surgery, psychiatry and psychology, allied health professionals, pediatrics, general practice, palliative care, acute care and unspecified.
*This table excludes 23 clinicians i.e. those for whom the options were never displayed (n=20) and those who did not have any of the characteristics listed here in the dataset (n=23).*

Among 89,050 orders where an alternative was selected, the unit cost of the alternative medication was the same for most orders (n=41,212; 58.4%), while 18,629 (26.4%) had lower cost and 10,728 (15.2%) alternate orders had higher cost. Clinicians selected a different pharmacy (e.g. Walgreens, CVS, etc.) for 39,634 (44.5%) of the alternative orders and a different pharmacy type (mail vs retail) for 7,508 (12.7%) of the alternate medication orders. A slight majority of these pharmacy type switches were from retail to mail order pharmacies [n= 4,680 (62.3%)]. **(Table 4)**

**Table 4:** Characteristics of medications selected from RTPB tools at three academic medical centers from April 2019 to October 2021.

| Characteristic | Site A | Site B | Site C | Total | p-value |
| --- | --- | --- | --- | --- | --- |
| **Total, n** | 17,596 | 1,429 | 70,025 | 89,050 |  |
| **Change in Total Cost** |  |  |  |  | <0.001 |
| Same alternative cost | 9018 (55.7%) | 366 (28.7%) | 34028 (64.1%) | 43412 (61.5%) |  |
| Lower Alternative cost | 3837 (23.7%) | 679 (53.2%) | 11293 (21.3%) | 15809 (22.4%) |  |
| Higher alternative cost | 3342 (20.6%) | 231 (18.1%) | 7775 (14.6%) | 11348 (16.1%) |  |
| Missing<sup>a</sup> | 1,399 | 153 | 16,929 | 18,481 |  |
| **Change in unit Cost** |  |  |  |  | <0.001 |
| Same cost | 7988 (49.3%) | 334 (26.2%) | 32890 (61.9%) | 41212 (58.4%) |  |
| Lower RTPB cost | 5689 (35.1%) | 693 (54.3%) | 12247 (23.1%) | 18629 (26.4%) |  |
| Higher RTPB cost | 2520 (15.6%) | 249 (19.5%) | 7959 (15.0%) | 10728 (15.2%) |  |
| Missing<sup>a</sup> | 1,399 | 153 | 16,929 | 18,481 |  |
| **Unit price change, mean (SD)** | 0.2 (3.0) | 1.2 (4.8) | 0.2 (5.3) | 0.2 (4.8) | <0.001 |
| **Total cost variance, mean (SD)** | 2.2 (46.7) | 35.4 (180.1) | 2.1 (82.1) | 2.7 (78.6) | <0.001 |
| **Change in quantity** |  |  |  |  | <0.001 |
| Same quantity | 11273 (64.1%) | 1082 (75.7%) | 54876 (78.4%) | 67231 (75.5%) |  |
| Lower RTPB quantity | 1744 (9.9%) | 120 (8.4%) | 6819 (9.7%) | 8683 (9.8%) |  |
| Higher RTPB quantity | 4579 (26.0%) | 227 (15.9%) | 8330 (11.9%) | 13136 (14.8%) |  |
| **Change in Pharmacy Type** |  |  |  |  | <0.001 |
| Same Pharmacy Type | 13,995 (80.0%) | 1370 (96.1%) | 36,170 (90.2%) | 51,535 (87.3%) |  |
| Different Pharmacy Type | 3,508 (20.0%) | 56 (3.9%) | 3,944 (9.8%) | 7,508 (12.7%) |  |
| Missing<sup>a</sup> | 93 | 3 | 29,911 | 30,007 |  |
| **Pharmacy Type Change Details** |  |  |  |  | <0.001 |
| Change: Mail Order to Retail | 1701 (9.7%) | 10 (0.7%) | 1117 (1.6%) | 2828 (3.2%) |  |
| Change: Retail to Mail Order | 1807 (10.3%) | 46 (3.2%) | 2827 (4.0%) | 4680 (5.3%) |  |
| Always Retail | 13,028 (74.0%) | 1,278 (89.4%) | 35,383 (50.5%) | 49,689 (55.8%) |  |
| Always Mail Order | 967 (5.5%) | 92 (6.4%) | 787 (1.1%) | 1,846 (2.1%) |  |
| Other<sup>b</sup> | 93 (0.5%) | 3 (0.1%) | 29911 (42.7%) | 30007 (33.7%) |  |
| **Change in Pharmacy** |  |  |  |  | <0.001 |
| Same Pharmacy | 8508 (48.4%) | 1342 (93.9%) | 39566 (56.5%) | 49416 (55.5%) |  |
| Different Pharmacy | 9088 (51.6%) | 87 (6.1%) | 30459 (43.5%) | 39634 (44.5%) |  |
| **Therapeutic Class** |  |  |  |  | <0.001 |
| Analgesics and Anesthetics | 1352 (7.7%) | 54 (3.8%) | 5830 (8.3%) | 7236 (8.1%) |  |
| Anti-infectives | 2116 (12.0%) | 204 (14.3%) | 5402 (7.7%) | 7722 (8.7%) |  |
| Cardiovascular | 3912 (22.2%) | 543 (38.0%) | 13015 (18.6%) | 17470 (19.6%) |  |
| Endocrine | 1570 (8.9%) | 138 (9.7%) | 6419 (9.2%) | 8127 (9.1%) |  |
| Gastrointestinal | 1419 (8.1%) | 95 (6.6%) | 4188 (6.0%) | 5702 (6.4%) |  |
| Hematological | 171 (1.0%) | 8 (0.6%) | 781 (1.1%) | 960 (1.1%) |  |
| Neurological | 747 (4.2%) | 52 (3.6%) | 14287 (20.4%) | 15086 (16.9%) |  |
| Psychiatric | 2124 (12.1%) | 107 (7.5%) | 0 (0.0%) | 2231 (2.5%) |  |
| Respiratory | 628 (3.6%) | 21 (1.5%) | 5171 (7.4%) | 5820 (6.5%) |  |
| Topical | 795 (4.5%) | 12 (0.8%) | 10855 (15.5%) | 11662 (13.1%) |  |
| Other <sup>c</sup> | 2762 (15.7%) | 195 (13.6%) | 4077 (5.8%) | 7034 (7.9%) |  |

<sup>a</sup> Missing values were not included for chi-square comparisons
<sup>b</sup> Others include specialty or unspecified pharmacy type
<sup>c</sup> Others include anti-neoplastic, biological agents, anti-allergic, diuretics, contraceptives, herbal and multivitamins.

In the adjusted analyses, the RTPB tool was more likely to display for older patients (risk ratio [RR]: 1.003; 95% CI: 1.002-1.003) relative to younger patients, and patients from sites A (RR: 1.423; 95% CI: 1.346-1.504) and C (RR: 1.967; 95% CI: 1.889-2.048) relative to site B. The RTPB tool was less likely to display for Black patients (RR: 0.940; 95% CI: 0.921-0.959) and Hispanic patients (RR: 0.874; 95% CI: 0.848-0.899) relative to White patients, and for patients with Medicaid (RR: 0.093; 95% CI: 0.088-0.099), Medicare (RR: 0.892; 95% CI: 0.874-0.909), and veteran/ military insurance (RR: 0.764; 95% CI: 0.733, 0.796) relative to commercial insurance. **(Table 5**)

**Table 5:**
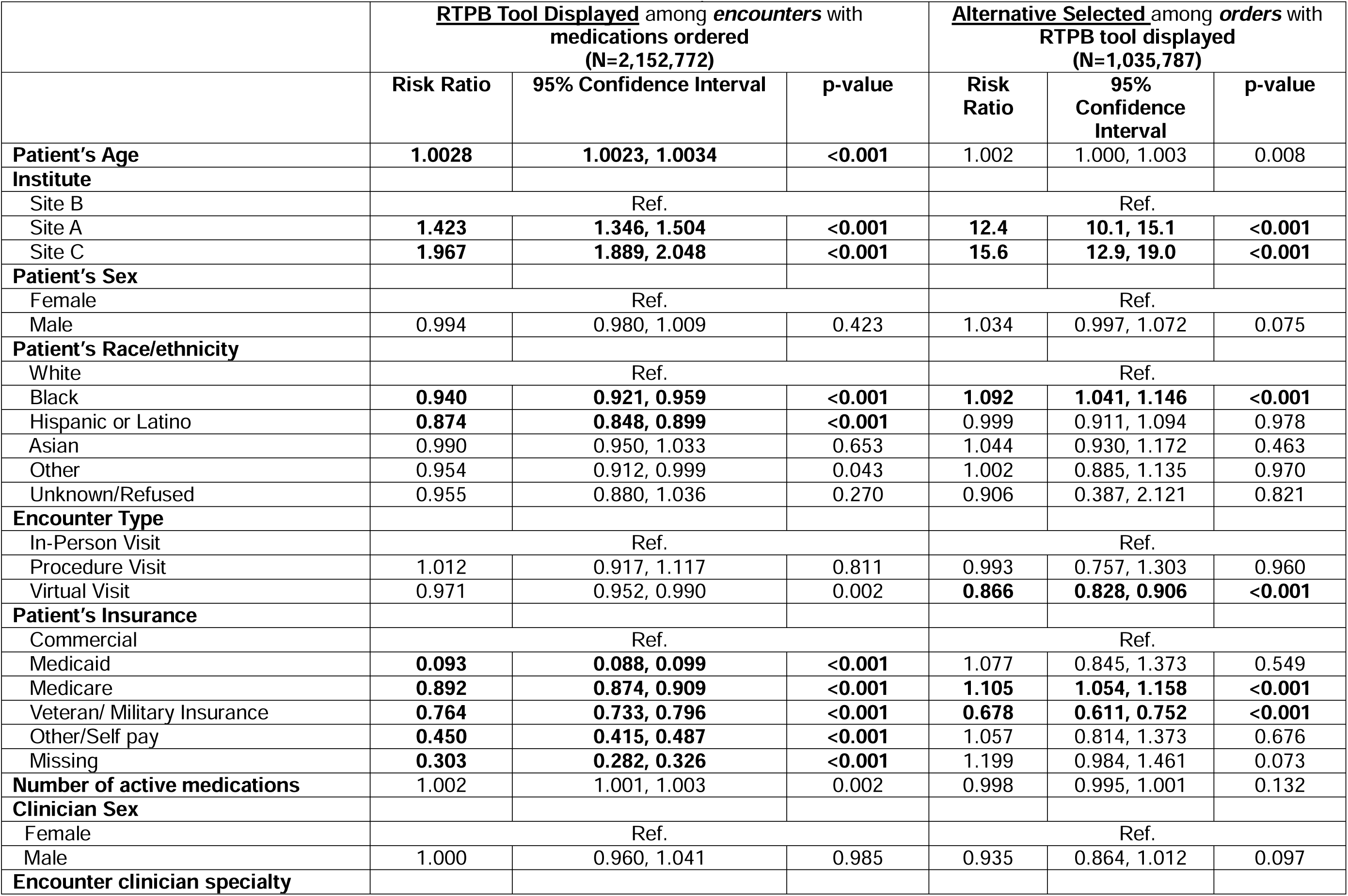
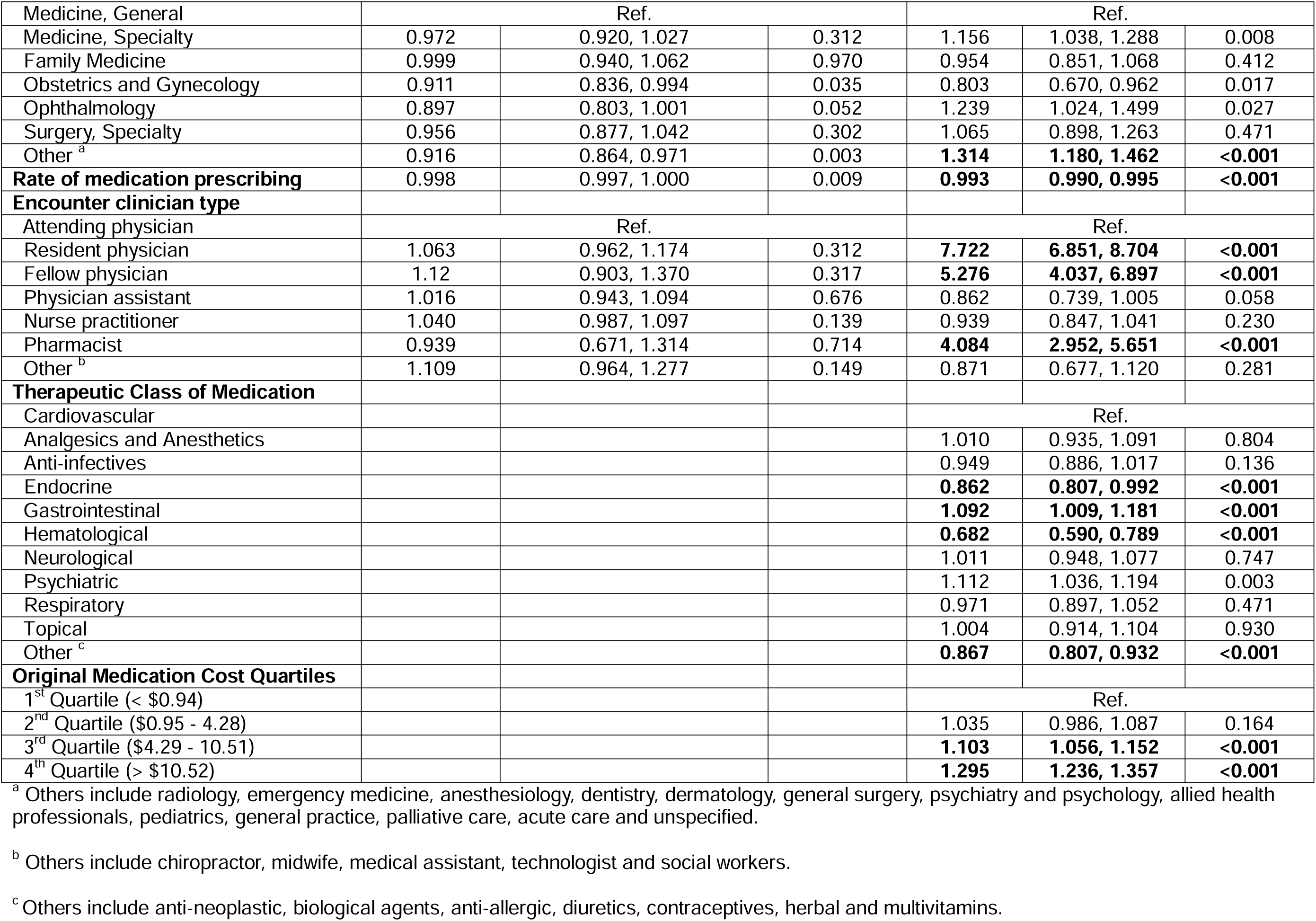
Multivariable regression analysis of patient and clinician characteristics associated with the display of the RTPB tool (encounter level data) and selection of alternative (order level data) at the three sites from April 2019 to October 2021.

Clinicians were more likely to select an alternative for patients at sites A (RR: 12.400; 95% CI: 10.100-15.100) and site C (RR: 15.6; 95% CI: 12.9, 19.0) relative to site B, and for Black patients (RR: 1.092; 95% CI: 1.041-1.146) relative to White patients. They were less likely to select an alternative for patients presenting for virtual visits (RR: 0.866; 95% CI: 0.828, 0.906). Clinicians were less likely to select an alternative for patients with veteran/ military insurance (RR: 0.678; 95% CI: 0.611- 0.752). In contrast to the display results, we found clinicians were more likely to select an alternative for those with Medicare (RR: 1.105; 95% CI: 1.054-1.158) relative to commercial insurance. Resident physician (RR: 7.722; 95% CI: 6.851, 8.704), fellow physician (RR: 5.276; 95% CI: 4.037, 6.897) and pharmacists (RR: 4.084; 95% CI: 2.952, 5.651) were more likely to select an alternative relative to attending physicians. Clinicians were less likely to select an alternative when prescribing endocrine (RR: 0.862; 95% CI: 0.807, 0.992) and hematological medications (RR: 0.682; 95% CI: 0.590, 0.789), and more likely to select an alternative when prescribing gastrointestinal medications (RR: 1.092; 95% CI: 1.009, 1.181) relative to cardiovascular medications. Clinicians were more likely to select an alternative for medications in the 3^rd^ cost quartile (RR: 1.103; 95% CI: 1.056, 1.152) and 4^th^ cost quartile (RR: 1.295; 95% CI: 1.236, 1.357) relative to medications within the 1^st^ cost quartile.

## DISCUSSION

Our study helps identify real-world evidence gaps in RTPB effects, finding that institutional implementation choices impacted utilization of the tool, including active vs passive alerting and cost thresholds. We also found that patient insurance, PBM coverage, and class of medication impacted display or selection of drug alternatives. Overall, though, the impact of the tool was relatively low, and less than previously reported studies.

### IMPLEMENTATION VARIANCES

RTPB tools differed in certain characteristics across the three sites including manually requesting RTPB estimates at site B, and contracting two RTPB vendors at site C. For the RTPB tools to be used during an encounter, the patient’s PBM must have a contract with the RTPB vendor. Having multiple RTPB vendors may therefore increase the likelihood that patient’s PBM is contracted. Resultantly we found that RTPB tools were least likely to display alternatives for clinicians at site B and more likely to display for clinicians at site C.

In addition, Site C had a higher prescription adjustment rate relative to the other two sites, which may be accounted for by including a cost differential threshold of >$3/month or >$0.10/day. In a similar study conducted at a single institution, Fitts et al reported a cost differential threshold of > $10 per fill or >$0.20/day, and that clinicians adjusted their prescriptions almost one-third of the time, which is three times higher than site C.(12) Therefore, no or low-cost differential display threshold may be a reason for low rates of prescription adjustment as the tool may have been unnecessarily triggered more frequently for medications that do not need to be changed resulting in alert fatigue. Adjusting the parameters of RTPB tools may therefore have an important role in influencing how clinicians interact with the tool.

### PATIENT CHARACTERISTICS

We found that even after adjusting for insurance types, RTPB tool was less likely to display for Black and Hispanic patients. It is possible that their particular insurance plan may not be compatible with the RTPB vendors and PBMs. We also found that the RTPB tool was less likely to display for patients with Medicare, Medicaid and Veteran/Military insurance relative to commercial insurance. This may expose the already vulnerable groups to high out of pocket medication costs and subsequently increased prescription non-fill rate. Bhardwaj et al. reported that display of RTPB tool was associated with a higher prescription fill rate (79.8% vs 71.7%) and lower cancelation rate (9.34% vs 14.89%).(8) Minoritized and underinsured groups are known to underutilize healthcare services.(13) Efforts are needed to avert the long-term consequences of racially and socioeconomically disparate use of RTPB tools.

### CLINICIAN CHARACTERISTICS

Among different clinician types, resident physicians, fellow physicians and pharmacists were more likely to select an alternative using the RTPB tool relative to attending physicians. Sloan et al. reported that younger primary care providers were more likely to interact with the RTPB tools at Duke University Health System.(14) Accreditation Council for Graduate Medical Education (ACGME) now mandates training about cost of care within residency programs, which might explain trainees’ and younger physicians’ enthusiasm for engaging with these tools and selecting therapeutic alternatives.(15)

### MEDICATION COST

Interestingly, our analysis revealed that in approximately 60% of the cases where prescriptions were changed, there was no corresponding change in OOPC for patients. In contrast, a study from the University of Colorado Health reported that there was no change in OOPC in 10% of the cases where an alternate was selected.(9) Our finding suggests that many prescription adjustments might have been driven by considerations other than cost including patient pharmacy preference. Almost half of the orders were changed to a different pharmacy (e.g. Walgreens, CVS, etc.) while one-tenth of the orders were changed to a different pharmacy type (mail vs. retail) among which a majority were switched from retail to mail order pharmacy.

Patients may prefer certain pharmacies or pharmacy types for a variety of reasons and convenience is an important consideration.(16) Patients may choose pharmacies that are closer to their home or workplace, offer extended hours, or provide home delivery services. In addition, mail-order pharmacies may be preferred by patients with chronic conditions requiring regular medication refills, as they can provide a more convenient and consistent supply without the need for frequent trips to a physical location. Patients may also have established trust and rapport with specific pharmacists or pharmacy staff, valuing personalized service and the pharmacist’s familiarity with their medication history.

### MEDICATION CLASSES

Alternative selection varied by medication class, and clinicians were less likely to select alternatives for endocrine and hematological medications, and more likely to select an alternative for gastrointestinal medications relative to cardiovascular medications. This variation could be influenced by several factors including the complexity of the diseases treated by these medications, the availability of suitable alternatives, and clinician familiarity and comfort with alternative options. For instance, the narrow therapeutic index of many endocrine and hematological medications may limit the substitution options, leading clinicians to adhere more strictly to prescribed treatments. In contrast, the broader range of efficacious and interchangeable medication options available for gastrointestinal conditions may encourage more frequent switching. Additionally, local formulary restrictions and insurance coverage might also play an important role in influencing the choice of alternatives, impacting how clinicians decide on the most suitable and cost-effective treatment options for their patients. Understanding these dynamics is crucial for optimizing tool design and educational efforts aimed at supporting clinician decision-making in selecting medication alternatives.

We found that clinicians were more likely to select an alternative for more expensive medications. Dessai et al reported that for high-cost drug classes, the use of RTPB tool reduced out-of-pocket costs by 40% relative to 11% for all drug classes overall.(7) Therefore, it may be beneficial, if alternatives suggested by the tool are regularly reviewed by interdisciplinary teams comprising of social workers, clinicians and pharmacy experts to ensure that alternatives are useful both clinically and financially. It could also be reasonable to consider RTPB implementation decisions to include a threshold for higher cost drug classes to maximize value and adoption while minimizing pop up burden.

This real-world study assessing the use of RTPB tools across multiple large US academic health systems is not without limitations. Academic centers may not generalize across the US, though all organizations had community footprints. The multicenter nature enhanced our understanding by incorporating diverse implementation characteristics of these tools. This quantitative analysis is limited in providing explanatory learnings, and qualitative research may further provide information for why certain behaviors were observed. Additional limitations include the retrospective nature of the study. While we only included encounter types that were eligible for an RTPB transaction, we are unable to determine the exact eligibility of specific orders, the role of pharmacy benefit manager coverage, cash-based alternative pricing, and impact of prior authorizations.

In summary our study found that RTPB tools were utilized at low rates which varied across the three institutions. Several patient sociodemographic, medication and clinician characteristics were associated with the use of the tool. RTPB design and implementation features such as price threshold and type of RTPB vendor likely affect usage. Future studies may include focus group discussion among clinicians who are high vs low utilizers for identification of barriers and facilitators to adoption of these tools by healthcare workers. Studies should also aim to understand patient perspectives on the usefulness of RTPB tools.

## Data Availability

All data produced in the present work are contained in the manuscript.

## References

1. Doshi JA, Li P, Pettit AR, Armstrong KA. Association of patient out-of-pocket costs with prescription abandonment and delay in fills of novel oral anticancer agents. 2018.

2. Sloan CE, Ubel PA. Patients want to talk about their out-of-pocket costs–can real-time benefit tools help? Journal of the American Geriatrics Society. 2023;71(5):1365.

3. Kullgren JT, Fendrick AM, editors. The price will be right—how to help patients and providers benefit from the new CMS transparency rule. JAMA Health Forum; 2021: American Medical Association.

4. Administration NAaR. Code of Federal Regulations 2024. Available from: https://www.ecfr.gov/current/title-42/chapter-IV/subchapter-B/part-423/subpart-C#p-423.128(d)(4).

5. Eric C. Naples E. What Does CMS’ Real-Time Benefit Tool Final Rule Mean for the Healthcare Industry? 2022 [May 16, 2024]. Available from: https://arrivehealth.com/what-is-a-real-time-benefit-tool-rtbt-and-what-does-cmss-new-rtbt-final-rule-mean-for-the-healthcare-industry/.

6. Wong R, Mehta T, Very B, Luo J, Feterik K, Crotty BH, et al. Where Do Real-Time Prescription Benefit Tools Fit in the Landscape of High US Prescription Medication Costs? A Narrative Review. Journal of general internal medicine. 2023;38(4):1038–45.

7. Desai SM, Chen AZ, Wang J, Chung W-Y, Stadelman J, Mahoney C, et al. Effects of real-time prescription benefit recommendations on patient out-of-pocket costs: a cluster randomized clinical trial. JAMA Internal Medicine. 2022;182(11):1129–37.

8. Bhardwaj S, Merrey JW, Bishop MA, Yeh H-C, Epstein JA. Impact of real-time benefit tools on patients’ access to medications: a retrospective cohort study. The American Journal of Medicine. 2022;135(11):1315–9. e2.

9. Sinaiko AD, Sloan CE, Soto MJ, Zhao O, Lin C-T, Goss FR. Clinician response to patient medication prices displayed in the electronic health record. JAMA Internal Medicine. 2023;183(10):1172–5.

10. Luo J, Wong R, Mehta T, Schwartz JI, Epstein JA, Smith E, et al., editors. Implementing real-time prescription benefit tools: Early experiences from 5 academic medical centers. Healthcare; 2023: Elsevier.

11. Follmann D, Proschan M, Leifer E. Multiple outputation: inference for complex clustered data by averaging analyses from independent data. Biometrics. 2003;59(2):420–9.

12. Fitts A, Teare AJ, Nelson SD. Price transparency at the point of prescribing with real-time prescription benefits. American Journal of Health-System Pharmacy. 2024:zxae108.

13. Scarborough JE, Bennett KM, Pietrobon R, Kuo PC, Pappas TN. Trends in the utilization of high-volume hospitals by minority and underinsured surgical patients. The American Surgeon. 2010;76(5):529–38.

14. Sloan CE, Morton-Oswald S, Smith VA, Sinaiko AD, Bowling CB, An J, et al. Real-world use of a medication out-of-pocket cost estimator in primary care one year after Medicare regulation. 2024.

15. Education ACfGM. Guide to the Common Program Requirements (Residency) 2024 [September 22, 2024]. Available from: https://www.acgme.org/globalassets/pdfs/guide-to-the-common-program-requirements-residency.pdf.

16. Patel PM, Vaidya V, Osundina F, Comoe DA. Determining patient preferences of community pharmacy attributes: a systematic review. Journal of the American Pharmacists Association. 2020;60(2):397–404.

